# Parsing clinical and neurobiological sources of heterogeneity in depression

**DOI:** 10.1101/2022.12.07.22283225

**Authors:** Kayla Hannon, Ty Easley, Wei Zhang, Daphne Lew, Aristeidis Sotiras, Yvette I. Sheline, Andre Marquand, Deanna M. Barch, Janine D. Bijsterbosch

**Author notes:** Corresponding authors: Kayla Hannon, Janine Bijsterbosch.

## Abstract

**Importance:** Patients with depression vary from one-another in their clinical and neurobiological presentation, yet the relationship between clinical and neurobiological sources of variation is poorly understood. Determining sources of heterogeneity in depression is important to gain insights into its diverse and complex neural etiology.

**Objective:** This study aims to determine how clinical and neurobiological sources of variation in depression interact to give rise to population heterogeneity. Specifically, we aimed to test if depression heterogeneity is characterized by subgroups that differ both clinically and neurobiologically and/or whether multiple neurobiological profiles give rise to the same clinical presentation.

**Design:** Clinically dissociated groups were selected to isolate clinical characteristics of depression (symptoms of anhedonia, depressed mood, and somatic disturbance; severity indices of lifetime chronicity and acute impairment; and late onset). Residual neurobiological heterogeneity within each group was assessed using neurobiologically driven clustering.

**Setting:** This study utilizes population-based data from the UK Biobank over multiple imaging sites.

**Participants:** All depressed participants (N=6,121) met one of the three clinical criteria: ICD10 label for depressive episode(s), probable MDD status, one or more recorded depression episodes. Control participants (N=8,565) reported minimal depression scores and no history of depression.

**Exposure:** There are no interventions or exposures in this study.

**Main Outcomes and Measures:** This study used several clinical features, multimodal MRI, and outcome phenotypes.

**Results:** The six clinically dissociated subgroups (total N=1909, n male=771, mean[SD] age=62.64[7.78]; subgroups: 211<n<576) had significantly larger (p<0.005, CI<[-0.260,-0.042]) neurobiological normative deviations than a comparison heterogeneous group (n=4210) and had distinct neurobiological profiles from each other. Imaging driven clustering within each clinically dissociated group identified two stable subtypes within the acute impairment group that differed significantly (p=0.003, CI:[-1.23,-0.345]) in cognitive ability, despite identical clinical profiles.

**Conclusions and Relevance:** The study identified distinct neurobiological profiles related to particular clinical depression features that may explain inconsistencies in the literature and sub-clusters within the acute impairment group with cognitive differences that were only differentiable by neurobiology. Our results provide evidence that multiple neurobiological profiles may give rise to the same clinical presentation, emphasizing the presence of complex interactions between clinical and neurobiological sources of heterogeneity.

**Study type:** Cross-sectional study

**Key Points:** *Question:* How do clinical and neurobiological sources of variation in depression interact to give rise to population heterogeneity?

*Findings:* In this cohort study, we identified statistically significant neurobiological profiles distinct to dissociable clinical features of depression and provide evidence for residual neurobiological heterogeneity after tightly controlling clinical variation, resulting in clusters with statistically significant differences in cognition.

*Meaning:* These results provide insights into the complex etiological relationships between neurobiological and clinical variation in depression and inform future biotype research by highlighting the need to distinguish biotypes both clinically and neurobiologically.

## Introduction

Depression is one of the most common mental health disorders with a lifetime prevalence of 27%^1^. Despite its prevalence, depression often goes underdiagnosed (especially in middle and older age^2^), and treatment efficacy is poor such that only 30% of patients respond to the first line of treatment^3^. Multiple neuroimaging correlates of depression have been identified, including structural, diffusion, and resting state abnormalities in the frontal cortex, deep nuclear gray matter, and temporal and parietal regions^4–6^. However, these findings have suffered from inconsistencies between reported results^5,7^ and/or small magnitude of effect^8^. Several prominent studies have suggested that such inconsistencies are largely due to heterogeneity (i.e., variability among individuals in their clinical symptomatology and/or neurobiology)^5,7,9–11^. The goal of this work is to parse the heterogeneity of depression to determine more robust neuroimaging correlates and gain insight into depression etiology.

Prior work to investigate heterogeneity in depression includes initiatives such as RDoC and HiTOP, which have defined transdiagnostic clinical commonalities such as RDoC’s Loss subconstruct^12^ and HiTOP’s Internalizing disorders^13^. Data-driven approaches have performed clustering on depression symptom scores to identify clinical subtypes^14–19^. Alternatively, other studies have focused on neurobiological sources of heterogeneity by performing data-driven clustering analyses on neuroimaging biomarkers^20,21^, albeit with challenges linked to overfitting^22^. Despite the valuable insights gained in these studies, the lack of convergence in observed depression subtypes across the emerging literature may suggest that the nature of depression heterogeneity is more complex and multifaceted than previously considered.

Heterogeneity is often implicitly or explicitly conceptualized as the presence of discrete subtypes within the overall population. Each subtype is thought to be characterized by a specific set of clinical features which arise from a specific set of neurobiological features, which we refer to as a one-to-one brain-symptom mapping (Fig. 1A). In this conceptualization, the same subtypes could be derived by investigating either the neurobiological or clinical features in isolation. Alternatively, it is possible that a specific set of clinical features may arise from multiple different neurobiological patterns in different individuals, which we refer to as many-to-one brain-symptom mapping (Fig. 1B; previously described as equifinality^23^). Note that one-to-one mapping may be multivariate in nature, such that a set of symptoms may be linked to a complex neurobiological pattern. In this study, we develop a novel two-stage framework to test one-to-one and many-to-one theories of depression heterogeneity. Notably, one-to-one and many-to-one mappings are not mutually exclusive, some mappings between brain and symptoms can be one-to-one and others can be many-to-one.

**Figure 1.**
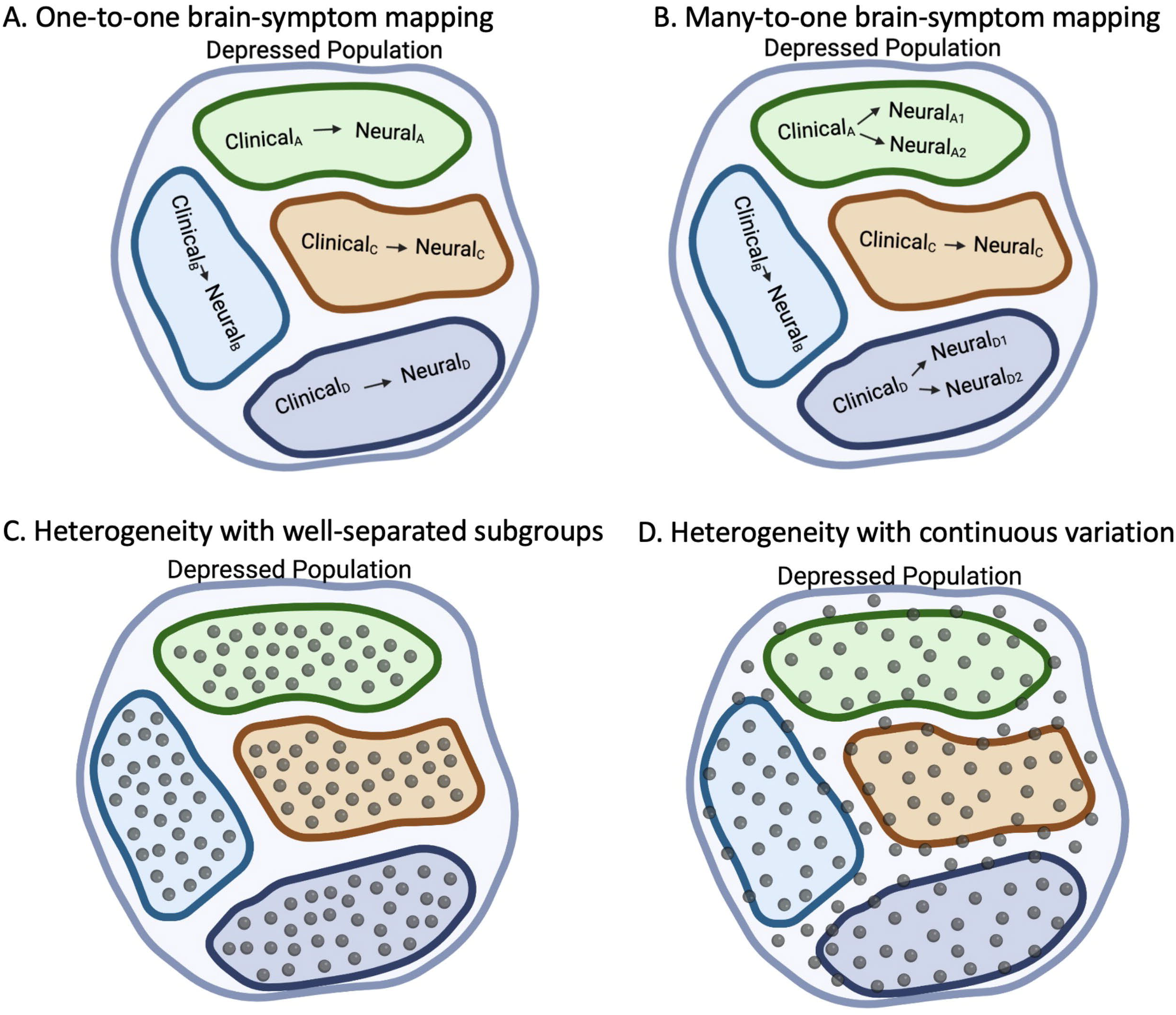
Theoretical perspectives on the relationship between clinical and neurobiological heterogeneity in the depression population. **(A)**. In the one-to-one theory, each clinical feature is expected to have a one-to-one association with a neurobiological explanation. **(B)**. In the many-to-one theory, multiple distinct neurobiological mechanisms can give rise to the same clinical presentation. Note that the one-to-one and many-to-one theories are not mutually exclusive and many-to-one mapping may be observed in some clinically dissociated groups, but not others. Heterogeneity may either take the form ofwell-separated clusters **(C)** or may vary along more continuous dimensions **(D)**. For example, prior evidence consistently finds distributions are more continuous in nature (e.g., see Fig. 1f in Drysdale et al^21^ and Fig. 3B in Wen et al^23^).

**Figure 2.**
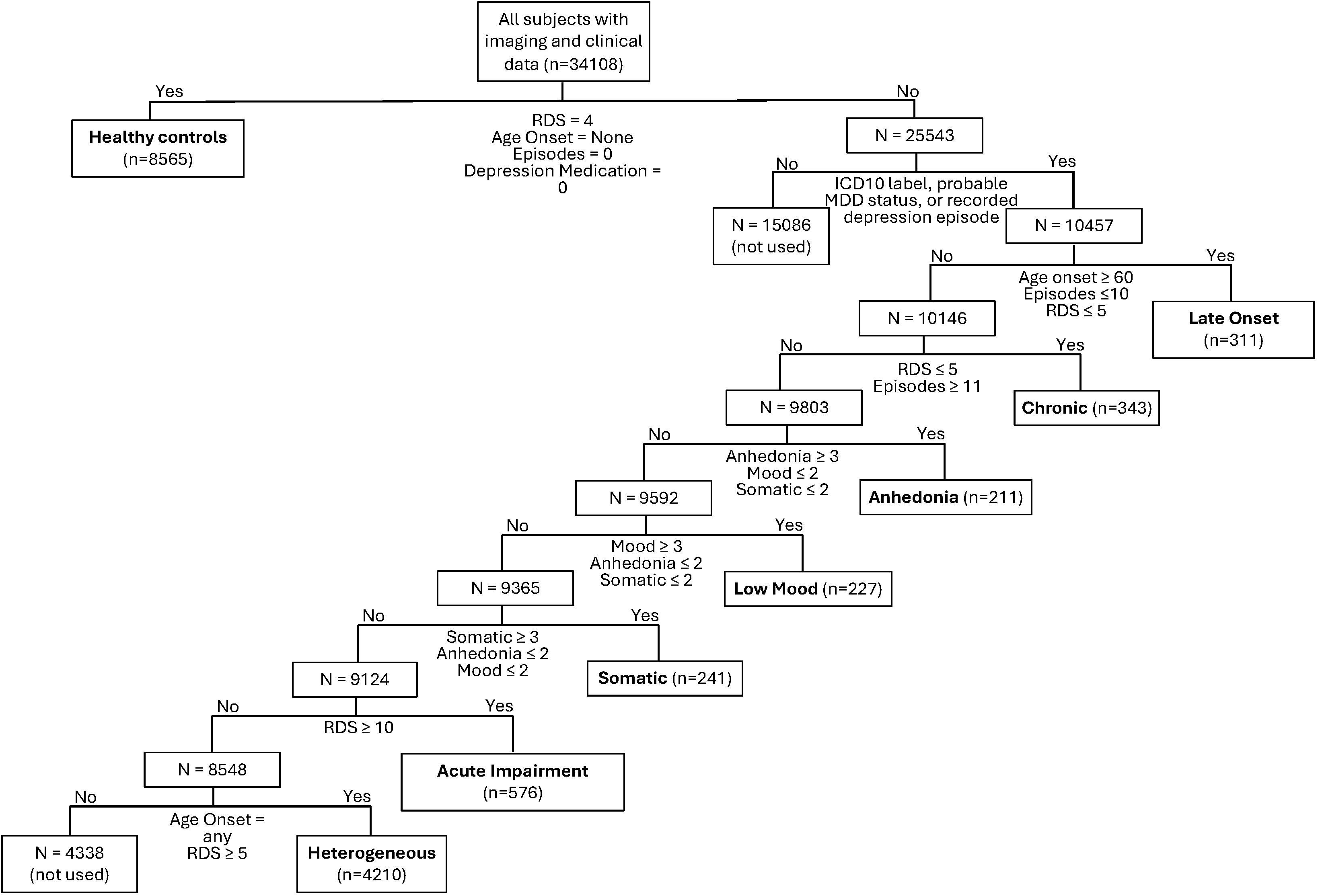
Clinically Dissociated Group Information. Inclusion criteria for clinically dissociated group. All criteria need to be met to be sorted into a group. For example, individuals in the anhedonia group scored high in anhedonia but low in depressed mood and somatic disturbance, were not late onset (first episode of depression at 60 years or older) nor had a high number of reported depressive episodes in their lifetime. The symptom-based clinically dissociated groups and the acute impairment group had criteria overlap but no participant overlap because the symptom groups were extractedfirst and therefore not available for the acute impairment group. The late onset group was comprised of individuals who experienced theirfirst episode of depression at 60 years or older but were not currently experiencing high symptoms nor had more than 10 episodes in their lifetime.

The goal of this study is to test one-to-one and many-to-one theories of depression heterogeneity by disentangling clinical and neurobiological sources of heterogeneity in depression. Specifically, we dissociate clinical features of depression that are typically highly collinear, which enables the investigation of potentially distinct neurobiological correlates of distinct clinical facets that jointly comprise depression. Based on prior suggestions that heterogeneity reduces effect sizes, we hypothesize that normative deviation magnitudes will be larger in resulting clinically dissociated groups relative to a heterogeneous comparison group. We furthermore hypothesized that clinically dissociated groups would be characterized by distinct neuroimaging profiles, in line with the one-to-one theory of depression heterogeneity. We then parse residual neurobiological heterogeneity within groups to investigate the presence of many-to-one mapping. We hypothesized that some clinically dissociated groups may contain stable and clinically relevant neurobiological subclusters (despite tightly controlling clinical heterogeneity), in line with the many-to-one theory of depression heterogeneity.

## Methods

### Dataset

This study utilizes data from the UK Biobank^25–27^. Depressed individuals were selected if they met at least one of three diagnostic criteria: i) an ICD10 label for F32 (depressive episode) or F33 (recurrent depressive episode; UKB variable ID 41270), and/or ii) probable MDD status as defined by ^28^ (UKB variable ID 20124 | 20125), and/ or iii) one or more recorded episodes of depression (UKB variable ID 4598 | 4631). Participants were placed in one of six dissociated clinical groups (anhedonia, low mood, somatic, chronic, late onset, acute impairment) or in the heterogeneous comparison group based on their self-response on the Recent Depressive Symptoms questionnaire (RDS)^8^, age of onset, and number of episodes (see Supplementary Methods, Table S1, and Table S2). Notably, the clinically dissociated groups were explicitly chosen to isolate clinical features along three axes of clinical variation (symptomatology, severity, age of onset), which enables the investigation of differential neurobiological profiles. These clinically dissociated groups were not intended to comprehensively map all individual variation in depression. Indeed, most individuals with depression present a multifaceted (rather than isolated) clinical profile and are grouped in the ‘heterogeneous group’, which is therefore representative of traditional depression study cohorts. Demographic information for the clinically dissociated groups is reported in Table S3.

### Imaging features

We investigated 90 multimodal imaging features that have previously been associated with depression in an older cohort^6^, especially in relation to the clinical features of interest (see Supplementary methods). For each imaging feature, normative modeling was used to estimate the normative deviation for all participants in the combined depressed groups (N=6,121) relative to healthy controls (N=8,565; see Supplementary methods for details and Fig. S1 for normative modeling accuracy measures).

### Statistical comparison of normative deviations across clinically dissociated groups

If heterogeneity led to decreased effect sizes in previous work, then clinically dissociated groups should be characterized by stronger neuroimaging normative deviations compared to a heterogeneous comparison group. To determine the imaging features that were significantly different between clinically dissociated groups, we applied two-sided one-way ANOVA with false discovery rate (FDR) multiple comparison correction over 90 imaging features. A two-sided t-test against 0 was performed for each imaging feature for each clinically dissociated group to assess their individual (rather than comparative) imaging profiles.

### ANOVA stability tests

We assessed the stability of the significant group difference replicating this analysis in a modest held-out dataset collected from UK Biobank participants released since the completion of the main analyses. Furthermore, we developed a null distribution of ANOVA F-statistics by randomly permuting participant labels (1000 permutations). To test for the impact of the larger heterogeneous group sample size, we subsampled the heterogeneous group down to N=300 (average sample size across the clinically dissociated groups) across 1000 bootstraps. Follow up details can be found in the Supplement.

### Data-driven clustering within clinically dissociated groups

Within each clinically dissociated group, clustering based on the 90 imaging features was performed to test for the possibility of many-to-one mapping (Fig. 1B). We did not perform clustering in the heterogeneous group because the goal of the clustering analysis was to assess the presence of residual neurobiological heterogeneity in clinically dissociated groups. The data-driven clustering pipeline comprised of feature reduction, cluster number estimation, cluster stability testing, and null comparison (see Supplementary methods for details).

### Cluster differentiation

To determine if the clusters were clinically valuable, we investigated whether any clusters significantly differed on the following depression-relevant phenotypes: cognitive ability (quantified using the PCA weights calculated in ^29^), Neuroticism (see Table S2), and Townsend socioeconomic deprivation index. We conducted unpaired t-tests between the clusters within each clinically dissociated group, using false discovery rate correction for the 6 comparisons. Cognitive ability was selected as it is associated with depression in older adults ^30^. Neuroticism was selected as it is a strong correlate of depression, indicative of “trait-like” depression^31^. Socioeconomic deprivation was chosen as it is an important modulator of depression^32^.

## Results

### Parse clinical heterogeneity

First we validated the clinically dissociated groups by assessing their longitudinal stability (F(5,2108)=32.14, p<0.008; Fig S2) and by verifying the groups differed in other depression related scores like the Generalized Anxiety Disorder and Neuroticism questionnaires (F(6,6120)=46.6, *p*_*FDR*_=4.2e-120; F(6,6120)=99.8, *p*_*FDR*_=3.6e-56 respectively; Fig. S3 and Table S4). To determine what imaging features the combined depressed group differs from controls on, a t-test against 0 was performed for all 90 imaging normative deviations (Fig. S4).

We hypothesized that the clinically dissociated groups are characterized by stronger neuroimaging normative deviations compared to a heterogeneous comparison group, in line with the one-to-one theory of heterogeneity and in support of the suggestion that heterogeneity leads to decreased effect sizes in the literature. ANOVA-based group comparisons revealed 10 out of 90 imaging features that showed significant group differences after correction for multiple comparisons (*F*(6,6212)>3.97 *p*_FDR_<0.0046; Fig. 3A-C). Tukey’s post-hoc analysis revealed that effects were driven predominantly by differences between the somatic, late onset, acute impairment, and heterogenous groups (Table S5). All ANOVA results achieved significance compared to a null distribution derived from random groupings (Fig. S5) and replicated in a modest held-out sample (Fig. S6; held-out demographics Table S6). Notably, the deviations for the heterogeneous group were significantly smaller (closer to zero in magnitude of deviation from controls) compared to at least one clinically dissociated group in all features, which was robust against subsampling to control for sample size effects (Fig. S7). These findings support the suggestion that heterogeneity contributed to reduced effect sizes in previous studies.

**Figure 3.**
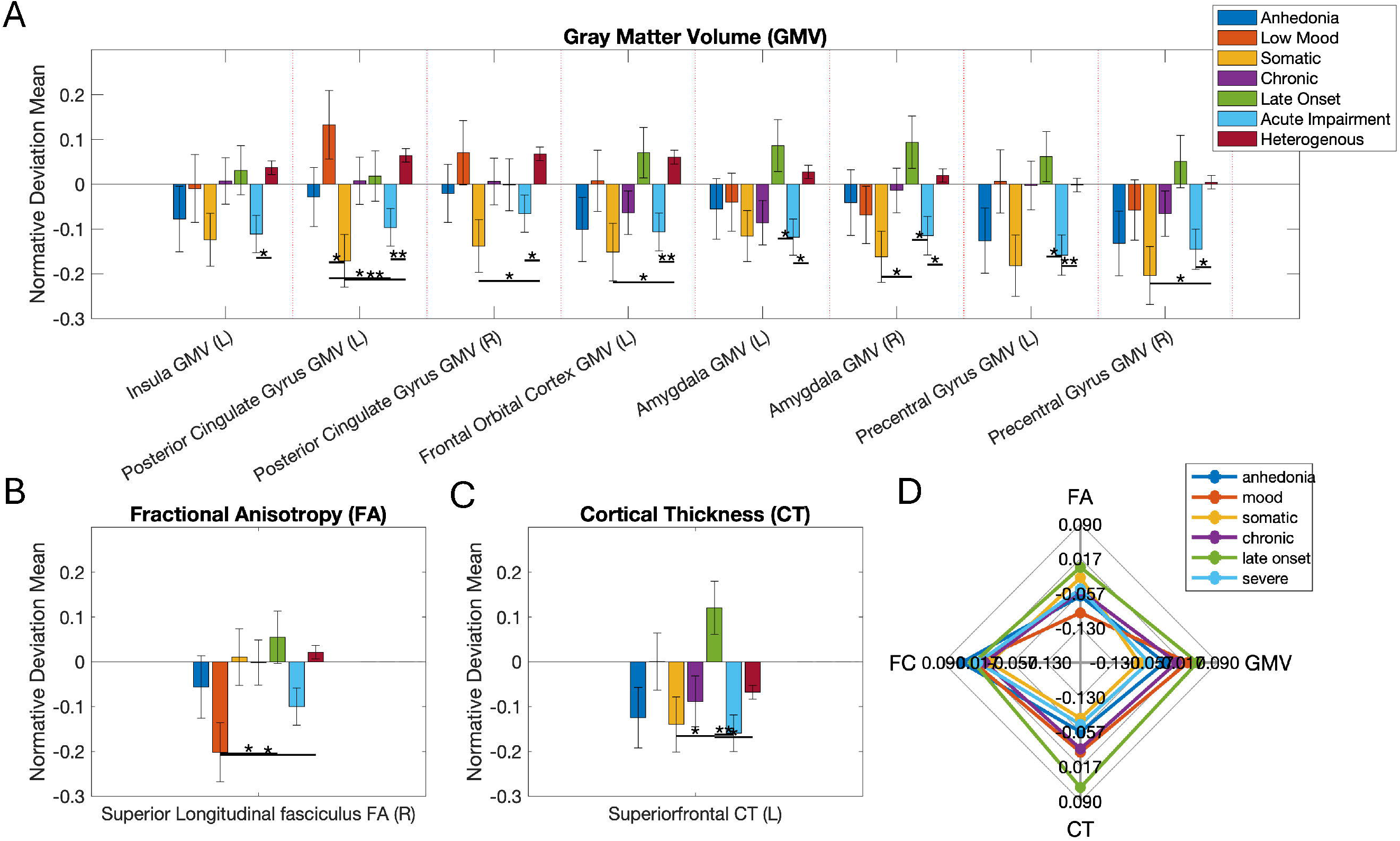
Distinct neurobiological profiles of clinically dissociated groups. **(A-C)** Results from an ANO VA to test which imaging measures differed significantly between 7 groups (6 clinically dissociated groups and the heterogeneous group). Bars show the group mean and error bars indicate standard error of mean. Results were significant afterfalse discovery rate multiple comparisons correction across the 90 imaging measures **A** is the gray matter volume results, B is the fractional anisotropy results, and C is the cortical thickness results. **(D)** Summary multimodal imaging profiles of the clinically dissociated groups, as represented by their mean normative deviations (relative to a healthy control population). FA = fractional anisotropy; GMV = gray matter volume; CT= cortical thickness. *signifies significance at p<0.05, **signifies p<0.01, and*** signifies p<0.001.

Investigating the group difference results in greater granularity. The acute impairment group differed from at least one other group with more negative normative deviations in most imaging features (superiorfrontal CT and insula, amygdala, posterior cingulate gyrus, precentral gyrus, and frontal orbital GMV). The late onset group differed from at least one other group in four imaging features (Fig. 3A-B) with more positive normative deviations than all the other groups in the amygdala and precentral gyrus GMV. The low mood group differed significantly from at least one other group in two imaging features, including positive normative deviations in posterior cingulate gyrus GMV. The somatic group differed significantly from at least one other group in six imaging features, including the precentral cortex, PCC, frontal orbital cortex, and amygdala. These findings reveal distinct neurobiological correlates of distinct clinical facets that jointly comprise depression.

### Neurobiological heterogeneity

We next investigated the potential presence of the many-to-one theory by assessing residual neurobiological heterogeneity within clinically dissociated groups. Our findings revealed that a 2-cluster solution was the optimal k in most bootstraps (cluster imaging profiles in Fig. S8; bootstraps in Fig. S9). The presence of stable participant clusters within clinically dissociated groups suggests the presence of some residual neurobiological variation even within groups that are highly clinically dissociated.

On visual inspection, the clusters were relatively continuous in nature (consistent with previous work^22,24^) and it is therefore important to determine the stability of the cluster assignments^22^. ARIs were above 0.6 for all clinically dissociated groups except for the low mood group, and all ARIs exceeded a null calculated from permuted data (Fig. 4A).

**Figure 4.**
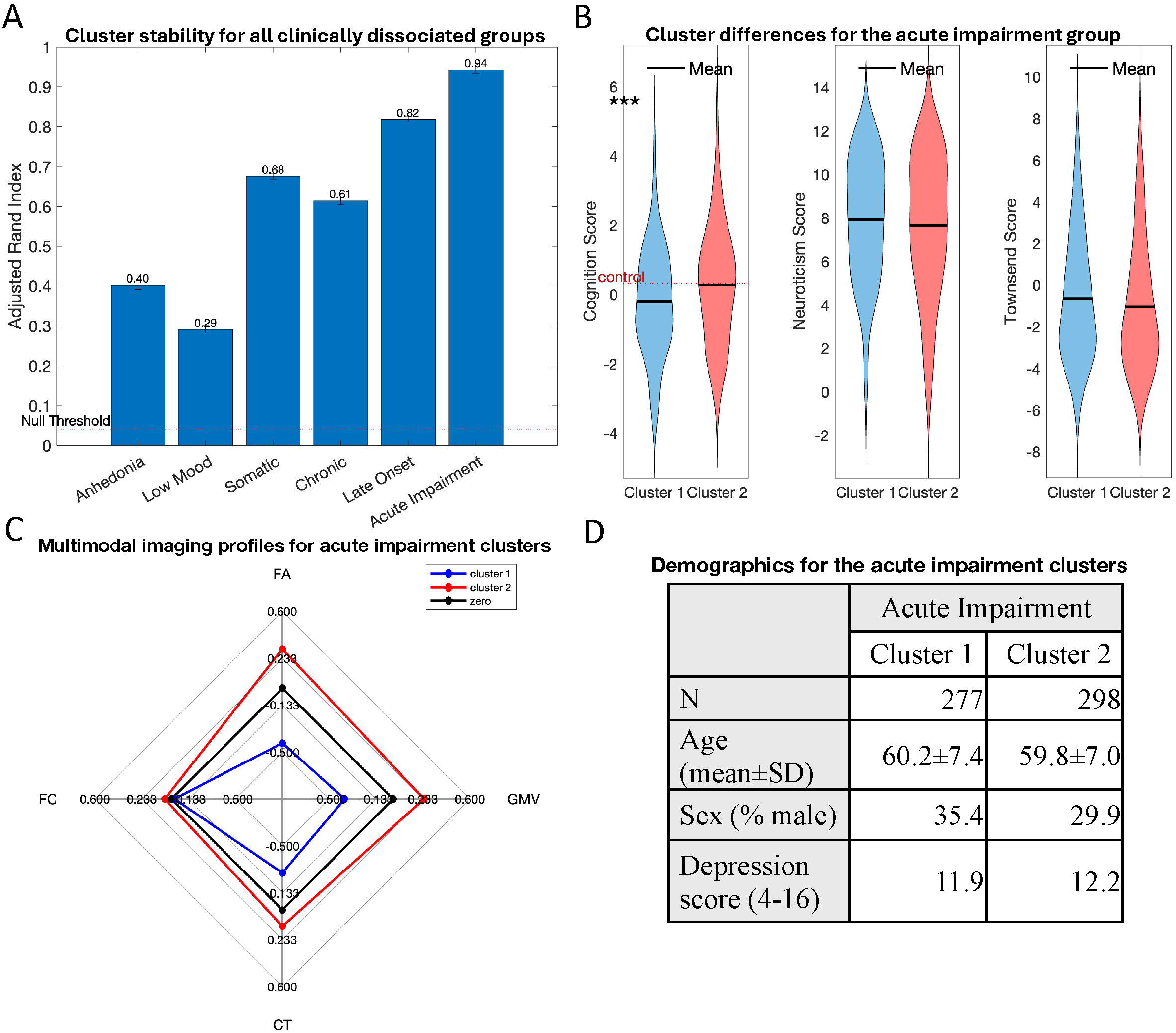
Stable and reproducible data-driven clusters within a clinically dissociated group. **(A)**. The Adjusted Rand Indices, which indicates similarity in clustering results across cross-validation folds adjustedfor random chance, for the clusters of each clinically dissociated group. The null threshold (95% confidence interval = (−0.014,0.087), mean = 0.016) was obtainedfrom permuted data, and error bars indicate the standard error of mean for each group. **(B)**. The clusters were further validated by investigating differences between clusters in cognitive ability, Neuroticism, and Townsend Deprivation Index. The mean cognitive score for the control group is the dashed red line. The clusters within the acute impairment groups differed significantly in cognition(*** p = 0.0034 postfalse discovery rate corrected). Cluster 1 is in blue, cluster 2 is in red, zero is in black. **(C)**. Summary multimodal imaging profiles of the acute impairment clusters. FA = fractional anisotropy; GMV = gray matter volume; CT = cortical thickness. **(D)**. Demographics of the acute impairment clusters. SD = standard deviation.

We furthermore tested the clinical utility of our data-driven clusters using depression-relevant phenotypes of cognitive score, trait neuroticism, and socioeconomic deprivation. The clusters in the acute impairment groups significantly differed in cognitive score after correction for multiple comparisons (*T(200)*=-3.5, *p*_FDR_=0.003; Fig. 4B & Table S7). In post hoc analysis, we determined specifically that the fluid intelligence measure (included in the composite cognition measure) significantly differed between the acute impairment clusters (*T(527)*=0.02, *p*_FDR_=0.004). The first cluster was characterized by reduced FA, GMV, and CT and had lower cognitive scores as compared to the second cluster, despite the clusters not differing significantly in age, sex, or depression score (p_FDR_>0.186). These findings are consistent with prior work^24^, and with hypothesized relationships between depression and cognition^33^. No significant cluster differences were observed for Neuroticism or the Townsend Deprivation Index post multiple comparisons correction (Table S7).

## Discussion

In this study we determined how clinical and neurobiological sources of variation in depression interact to give rise to the heterogeneity that is observed across the clinical population. Our results provide evidence to support the presence of both one-to-one and many-to-one heterogeneity in depression, emphasizing the presence of complex interactions between clinical and neurobiological sources of variation. Specifically, our findings revealed larger magnitude of neuroimaging normative deviations within clinically dissociated groups compared to a ‘typical’ heterogeneous comparison group, which is in line with the one-to-one theory of heterogeneity and points to the impact of heterogeneity in prior studies as an obfuscating factor. At the same time, we revealed evidence for residual neurobiological clusters that differed in cognitive ability within the acute impairment group, in line with the many-to-one theory of heterogeneity. To our knowledge, this is the first study to explicitly assess the presence of many-to-one mapping after tightly controlling for clinical heterogeneity, indicating that multiple distinct neurobiological mechanisms may give rise to the same clinical presentation for some clinical features. Moving beyond traditional subtyping approaches for depression heterogeneity, our novel hierarchical framework allowed us to dissociate neurobiologically distinct profiles for individual clinical features of depression and tease out residual neurobiological heterogeneity after tightly controlling clinical variation. Importantly, the resulting insights into the interactions between clinical and neurobiological sources of depression heterogeneity provide a strong foundation for future research into depression biotypes by highlighting the importance of concurrently modeling multiple axes of variation to distinguish biotypes based on both clinically and neurobiologically variables.

Our findings revealed that each isolated clinical depression feature was associated with distinct neuroimaging correlates (Fig. 3). Importantly, these results provide symptom-level insights into the neurobiological etiology of depression. Some clinically dissociated groups showed imaging feature profiles in line with prior findings, whereas other groups were characterized by unexpected imaging profiles. For example, our findings reveal reduced gray matter volume in the bilateral amygdala and posterior cingulate gyrus in the acutely impaired group and multiple symptom groups, consistent with prior work^34,35^. Furthermore, we showed that somatic disturbance was associated with reduced gray matter volume in the primary motor cortex, in line with previous research^36,37^. The late onset group, however, was unexpectedly characterized by positive normative deviations (i.e., preserved/increased gray matter volume relative to controls; Fig. 3A). This finding is in the opposite direction of prior work linking late onset depression with gray matter atrophy^38,39^, which is sometimes posited as inherent to late onset etiology^40^. However some research has shown that remittance in late onset depression was associated with relatively larger gray matter volume^41,42^. As our late onset group deliberately contains individuals who are currently not high in depression symptoms to achieve the desired dissociation of clinical features, our findings do not support the assertion that gray matter atrophy is inherent to late onset etiology, but are instead consistent with prior literature pointing to larger gray matter volume as a protective mechanism linked to remission^43^. Potential mechanisms for increased gray matter include protective neuroplasticity inducing effects of antidepressant medication^44^ with increased duration of exposure occurring during treatment of multiple depressive episodes. These findings showcase the value of dissociating clinical features to tease apart discrepancies in the literature and inform possible mechanisms.

More broadly, our results showed larger magnitudes of neuroimaging normative deviations in many of the clinically dissociated groups as compared to the comparison heterogeneous group, which supports the suggestion that heterogeneity is leading to smaller effect sizes^8–10^. In the age of population neuroimaging datasets, this finding points to the value of carefully parsing clinical heterogeneity to investigate one-to-one brain-symptom mapping and identify robust neuroimaging correlates.

Moving beyond one-to-one brain-symptom mapping, our results revealed the presence of residual neurobiological heterogeneity within the acute impairment clinically disassociated group. Using a unique hierarchical approach, we found stable neuroimaging-driven participant clusters that differed on cognitive ability in the acute impairment group, which indicates the presence of many-to-one mapping for this clinical feature. Importantly, the resulting clusters differed in cognition (specifically fluid intelligence), which was not explained by other clinical features such as depression severity or symptoms, socioeconomic deprivation, age, or sex. Specifically, one cluster was characterized by relatively impaired neurobiology and impaired cognitive ability, whereas the second cluster was characterized by relatively protective imaging features and preserved cognitive ability (e.g., similar to healthy controls). As such, the latter cluster may represent a cognitively resilient group despite matched symptoms of depression. The relationship between depression and reduced cognitive ability is well-established^24,33,45^, but the impact of depression severity (aka acute impairment) on cognitive ability is unclear due to inconsistent findings^46,47^. Our work provides insight into the potential reason for these inconsistent findings.

The current study, while aided by rich data of the UK Biobank, had some limitations. We decided to tightly control the clinically dissociated groups to isolate individual clinical features, which was important to gain insights into sources of heterogeneity and to identify distinct neuroimaging patterns. However, the specific inclusions/exclusion criteria of our clinically dissociated groups focus on relatively atypical individuals because most individuals with depression have a more mixed clinical presentation. Future work may investigate the neuroimaging correlates of homogeneous groups with a more mixed clinical presentation to test whether the one-to-one brain-symptom mapping model extends to more complex and multivariate clinical profiles. The UKB does not perform structured clinical interview to confirm depression diagnosis, although we used alternative diagnostic criteria in line with prior work^8^. Nevertheless, follow-up work in clinical cohorts would be of interest. Additionally, this study focused on interactions between clinical and neurobiological heterogeneity, but future work may want to extend our approach into other sources of heterogeneity such as genetic variation^48^. Future efforts are also needed to further validate the neurobiological clusters identified in this work. The number of participants with complete data on clinically relevant phenotypes for differentiation was relatively small, which limited the number of phenotypes we could use for differentiation.

In summary, our results provide insights into the symptom-level neurobiological etiology of depression and provides evidence for both one-to-one and many-to-one models of heterogeneity. Our findings provide insights into distinct symptom-level neural correlates within the broader construct of depression that may explain discrepancies in the literature and shed light on possible mechanisms. Taken together, our results suggest the importance for future studies to account for clinical and neurobiological heterogeneity when trying to understand depression etiology and neurobiological mechanisms.

## Supporting information

UK Biobank Imaging Feature IDs

## Data Availability

UK Biobank data is available following an access application process. For more information: https://www.ukbiobank.ac.uk/enable-your-research/apply-for-access.

https://github.com/PersonomicsLab/Heterogeneity.

## Acknowledgements

This work is supported by the NIH (R01 MH128286).

## Data Sharing Statement

This research was performed under UK Biobank application number 47267. UK Biobank data (Miller et al. <u>2016</u>; Sudlow et al. <u>2015</u>) is available following an access application process. For more information: https://www.ukbiobank.ac.uk/enable-your-research/apply-for-access. All analysis code for this article is available at: https://github.com/PersonomicsLab/Heterogeneity.

