## Supplement for "Parsing clinical and neurobiological sources of heterogeneity in depression"

### Supplementary Methods

##### Imaging features

The imaging features investigated in this study were chosen as they have been previously associated with depression, particularly the clinical features investigated in this study (see Background Information on Clinically Dissociated Groups for more details). This literature driven approach avoids double dipping as the imaging features were not selected for their association with depression in the UK Biobank. These imaging features include select imaging derived phenotypes (IDPs) for gray matter volume, cortical thickness, white matter hyperintensity, fractional anisotropy, and functional networks (see Supplemental spreadsheet ‘UKB IDP IDs’). All imaging data were acquired on a 3T scanner. Gray matter volume and cortical thickness were obtained from T1-weighted images. White matter hyperintensity is from T2 fluid-attenuated inversion recovery (FLAIR) acquisition. Fractional anisotropy and functional networks are from diffusion MRI and functional MRI acquisition, respectively. Data processing for all neurobiological features has been described in previous work^1,2^.

##### Depression measure

Depression symptoms and acute impairment were measured based on responses to the RDS questionnaire (UKB variable IDs 2050, 2060, 2070, and 2080 using instance 2 data; Table S1). Age of onset was determined by report of first episode of depression via online questionnaire (UKB variable ID 20433, using instance 0 data; Table S1). Chronicity was defined by reported number of episodes on a touchscreen on day of scan (UKB variable ID 4620 and 5386, using instance 2 data; Table S1).

Supplementary Table S1. Clinical features used in study***.*** *The UK Biobank questions used to dissociate* *depression symptoms and other clinical features including the Recent Depressive Symptoms (RDS) measure used to quantify depression symptoms in this study.*

| Clinical Feature | Question | Answer Options |
| --- | --- | --- |
| RDS1 | ‘Frequency of depressed mood in last 2 weeks’ | (1) Not at all  (2) Several days  (3) More than half the days  (4) Nearly every day  Sum range: 4-16 |
| RDS2 | ‘Frequency of unenthusiasm/disinterest in last 2 weeks’ |  |
| RDS3 | ‘Frequency of tenseness/restlessness in last 2 weeks’ |  |
| RDS4 | ‘Frequency of tiredness/lethargy in last 2 weeks’ |  |
| Number of episodes | ‘How many periods have you had when you were feeling depressed or down for at least a whole week?' OR 'How many periods have you had when you were uninterested in things or unable to enjoy the things you used to for at least a whole week?' | Report number |
| Age of onset | ‘About how old were you the FIRST time you had a period of two weeks like this? (Whether or not you received any help for it.)’ | Report number |

##### Background information on clinically dissociated groups

Due to indications in the literature that they may exhibit distinct neuroimaging profiles, six clinical features were investigated: symptoms of anhedonia, depressed mood, and somatic disturbance; severity indices of lifetime chronicity and acute impairment; and late onset depression. Anhedonia, somatic disturbance, and depressed mood have consistently been found as symptom-based subtypes^1^. Anhedonia (inability to feel pleasure) has been linked to abnormality in Salience (SN) and Central Executive Networks (CEN)^7^ as well as decreased gray matter volume (GMV) of the amygdala^8,9^ and anterior cingulate gyrus (ACC)^10^. Somatic disturbance (increased or decreased motor activity) is associated with alterations in the precentral (primary motor) gyrus and anterior cingulate gyrus volumes^11,12^. Depressed mood (defined as low mood) is associated with reduced volume in frontal regions such as medial frontal cortex and ACC^13^. Severity is another important feature of depression, which takes two possible forms. Trait depression (i.e., baseline predisposition) leads to numerous episodes of depression throughout the lifetime which we refer to as chronic severity or chronicity. On the other hand, acute severity refers to state depression (being in a current state of severe depression). High chronicity has been associated with a reduction in hippocampal volume^14,15^. Acute severity, on the other hand, may be associated with more state-dependent changes such as abnormal functional connectivity, particularly in Default Mode Network (DMN)^16^. The final clinical feature relates to age of first depression episode onset. When depression episodes begin in late life (60 years or older) it is referred to as late onset. Late onset patients are more likely to have reduced fractional anisotropy (FA)^17^ and increased white matter hyperintensities (WMH)^18^ even after accounting for age. We decided to include the late onset group as the UK Biobank cohort skews older. The imaging features evaluated were selected to include features relevant to the clinically dissociated groups. Notably, all clinical features of symptoms, severity, and age of onset are highly collinear in typical depression cohorts. To our knowledge, our study is the first to select participants in order to dissociate these clinical features of depression and allow robust investigation into potentially distinct neuroimaging correlates. The prior neuroimaging findings summarized above were used to guide the selection of our imaging features.

##### Validation of clinically dissociated groups

We performed an ANOVA with 7 levels representing the six clinically dissociated groups and heterogeneous comparison group, to test whether groups differed on clinical scores of depression (using RDS and the Neuroticism Questionnaire; N) and anxiety (using the Generalized Anxiety Disorder questionnaire; GAD). The GAD was obtained at a separate timepoint (not on the day of scanning) via an online questionnaire (Table S2). Furthermore, we assessed the longitudinal stability of symptom scores (including the anhedonia, depressed mood, and somatic disturbance scores) using a previous data instance (on average 6 years before the instance investigated in the main analysis) and a follow up instance (on average 2 years after the instance investigated in the main analysis). Three separate ANOVAs were performed for anhedonia, mood, and somatic scores with a factor for instance (3 levels) and a factor for clinical group (6 levels).

##### Quality control of imaging data

The UK Biobank performs rigorous quality control for artifacts in T1-weighted images, using a machine learning algorithm trained on 5816 participants to flag potentially problematic datasets for manual inspection (including problematic head motion, signal to noise ratio, brain asymmetry, tissue outside of the brain mask, and much more). Datasets that did not pass quality control were not released. Additionally, resting state preprocessing included ICA-FIX to remove noise components, and diffusion preprocessing included eddy current correction (see Alfaro-Almagro et al^2^ for further details). The UK Biobank has relatively small site effects^2^ compared to other multi-site studies because UKB sites were purpose-built and used identical scanners and sequences. Nevertheless, we controlled for site effects by including site as a covariate in the normative model and have confirmed the clinically dissociated groups did not differ by scanner site (p = .341).

##### Normative modeling

To be highly sensitive to biologically meaningful variation, we employed normative modeling on the imaging features to acquire “normative deviations” that represent how each person differs from the healthy controls for each imaging measure, accounting for covariate effects. Normative modeling has successfully been applied to parse heterogeneity in other mental disorders^19–21^, including depression^22^. The normative model applied was SinhArcsinh-warped Bayesian linear regression^23^ with covariates age (UKB variable ID 21003, instance 2), sex (UKB variable ID 31, instance 2), head motion (UKB variable ID 25741, instance 2), and image acquisition site (UKB variable ID 54, instance 2). This algorithm was selected as previous research found it represented neuroimaging features most accurately of all algorithms assessed^23^. More precisely, the normative model was a b-spline basis expansion over the covariates (age, sex, head motion, acquisition site) with equally spaced knot points employing a SinhArcsinh warp to model non-linearity in the imaging feature (the response variable in this model). The prior weight precision, noise precision, and warp shape parameters were controlled by model hyper parameters which were estimated using an Empirical Bayes approach. The normative deviation is a Z-statistic of the model’s residuals, measuring how much the individual’s imaging feature deviated from the model’s expected value based on their covariates. These z-scores were calculated by subtracting the true imaging value from the estimated prediction (after warping), divided by the standard deviation accounting for all estimated variance components, as described in previous publications^24^.

##### Data-driven clustering steps:

- Feature reduction was performed using principal component analysis (PCA) applied to 90 imaging features keeping the minimum number of principal components that explain at least 75% of the variance as features in subsequent k-means clustering. This procedure and the below steps were performed independently within each clinically dissociated group.
- The optimal number of clusters was determined at each iteration. We measured the silhouette score for *k* from 2 through 10 to determine how many clusters is best. We bootstrapped this data 1000 times (randomly sampling the data with replacement) and determined optimal cluster number as the *k* that has the highest silhouette score the most times over the 1000 bootstraps. We performed the PCA, feature selection, and k-means clustering pipeline for each bootstrap. The feature selection (selecting the components that explained 75% of the variance) was highly consistent: the first 32 components explained 75% of the variance for every bootstrap. We also calculated the Calinski-Harabasz index, another cluster criterion, alongside the silhouette index.
- The stability of the clusters was determined. To do this, we performed 10-fold cross validation, which was repeated over 100 iterations. In each fold, 90% of the participants were used to repeat the PCA, selection of PCA components (33 components), and k-means clustering (k=2) at the found dimensionality. We calculated the Adjusted Rand Index (ARI) between each fold, which provides a measure of the stability of the cluster assignments. For each fold, we trained the PCA and k-means on 90% of data, then applied the PCA coefficients and k-means centroids to the remaining 10% culminating in cluster assignments for all 100% of data for that fold. In that iteration, the ARIs between cluster assignments for all 10 folds were computed. The mean ARI for that entire iteration was then calculated. This process was repeated for 100 iterations (with new folds every time) and the mean ARIs for each iteration were averaged (and STD calculated) one final time to get the final mean ARI and STD shown in Fig. 4A.
- Stability results were compared against a null model. We synthesized a null dataset by randomly permuting the participant labels 1000 times independently within each imaging feature (i.e., breaking the correlation between imaging features) within each clinically dissociated group. The resulting null data were processed through the pipeline described above (including PCA, component selection, and cross-validation performed separately within each clinically dissociated group), which resulted in a set of ARI thresholds based on null data.

### UK Biobank variable IDs

##### Supplementary Table S2 UK Biobank IDs

|  | **Imaging Features (Imaging Derived Phenotypes)**  UK Biobank IDs are on Excel sheet ‘UKB IDP IDs’ |
| --- | --- |
| **UKB ID** | **Clinically Dissociated** **Group Criteria** |
| 2050 | RDS 1 - Frequency of depressed mood in last 2 weeks |
| 2060 | RDS 2 - Frequency of unenthusiasm / disinterest in last 2 weeks |
| 2070 | RDS 3 - Frequency of tenseness / restlessness in last 2 weeks |
| 2080 | RDS 4 - Frequency of tiredness / lethargy in last 2 weeks |
| 4620 | Number of depression episodes |
| 5386 | Number of unenthusiastic/disinterested episodes |
| 20433 | Age at first episode of depression |
| **UKB ID** | **Anxiety measure (used for clinical comparisons between groups)** |
| 20506 | GAD 1 - Feeling nervous, anxious, or on edge |
| 20509 | GAD 2 - Not being able to stop or control worrying |
| 20520 | GAD 3 - Worrying too much about different things |
| 20515 | GAD 4 - Trouble relaxing |
| 20516 | GAD 5 - Being so restless that it is hard to sit still |
| 20505 | GAD 6 - Becoming easily annoyed or irritable |
| 20512 | GAD 7 - Feeling afraid as if something awful might happen |
| **UKB ID** | **Normative Modeling Covariates** |
| 21003 | Age when attended assessment centre |
| 31 | Sex |
| 25741 | Mean rfMRI head motion, averaged across space and time points |
| 54 | UK Biobank assessment centre |
| **UKB ID** | **Cluster Differentiation Phenotypes** |
| 20016 | Cognition 1 - Fluid intelligence score |
| 20023 | Cognition 2 - Mean time to correctly identify matches |
| 189 | Townsend deprivation index at recruitment |
| 1920 | Neuroticism 1 - Suffer from 'nerves' |
| 1930 | Neuroticism 2 - Miserableness |
| 1940 | Neuroticism 3 - Irritability |
| 1950 | Neuroticism 4 - Sensitivity / hurt feelings |
| 1960 | Neuroticism 5 - Fed-up feelings |
| 1970 | Neuroticism 6 - Nervous feelings |
| 1980 | Neuroticism 7 - Worrier / anxious feelings |
| 1990 | Neuroticism 8 - Tense / 'highly strung' |
| 2000 | Neuroticism 9 - Worry too long after embarrassment |
| 2010 | Neuroticism 10 - Suffer from 'nerves' |
| 2020 | Neuroticism 11 - Loneliness, isolation |
| 2030 | Neuroticism 12 - Guilty feelings |

#### Descriptive statistics of group differences in clinical scores

##### Supplementary Table S3. Demographic and Clinical Characteristics.

Means if not otherwise stated. RDS = Recent Depressive Symptoms measure (sum range 4-16), each individual RDS question has a range 1-4.

|  | Sample size | Age (mean±SD) | Sex  (% male) | RDS Sum | RDS Anhedonia | RDS Mood | RDS Restless | RDS Lethargy | Age Onset | Episode Number |
| --- | --- | --- | --- | --- | --- | --- | --- | --- | --- | --- |
| Anhedonia | 211 | 61.9±7.4 | 40.8 | 8.7 | 3.3 | 1.5 | 1.4 | 2.4 | 34.8 | 2.1 |
| Low Mood | 227 | 61.2±7.4 | 38.3 | 8.9 | 1.7 | 3.2 | 1.6 | 2.4 | 30.2 | 2.4 |
| Somatic | 241 | 63.6±7.5 | 41.9 | 8.8 | 1.5 | 1.5 | 3.3 | 2.5 | 31.0 | 2.7 |
| Chronic | 343 | 60.3±7.8 | 45.8 | 4.5 | 1.0 | 1.0 | 1.1 | 1.3 | 31.9 | 33.0 |
| Late Onset | 311 | 69.9±4.1 | 48.9 | 4.3 | 1.0 | 1.0 | 1.0 | 1.3 | 63.7 | 1.8 |
| Acute Impairment | 576 | 60.0±7.2 | 14.9 | 12.1 | 2.9 | 3.0 | 2.7 | 3.4 | 29.2 | 2.2 |
| Heterogenous | 4212 | 61.6±7.5 | 35.6 | 6.2 | 1.3 | 1.4 | 1.4 | 2.0 | 34.7 | 6.3 |
| Healthy Control | 8565 | 64.7±7.2 | 58.4 | 4.0 | 1.0 | 1.0 | 1.0 | 1.0 | NaN | 0.0 |

### Normative modeling accuracy

Supplementary Figure S1. Accuracy Measures for the Normative Models.

A normative model was run for each imaging feature. The Pearson correlation is between the true and predicted imaging feature values.

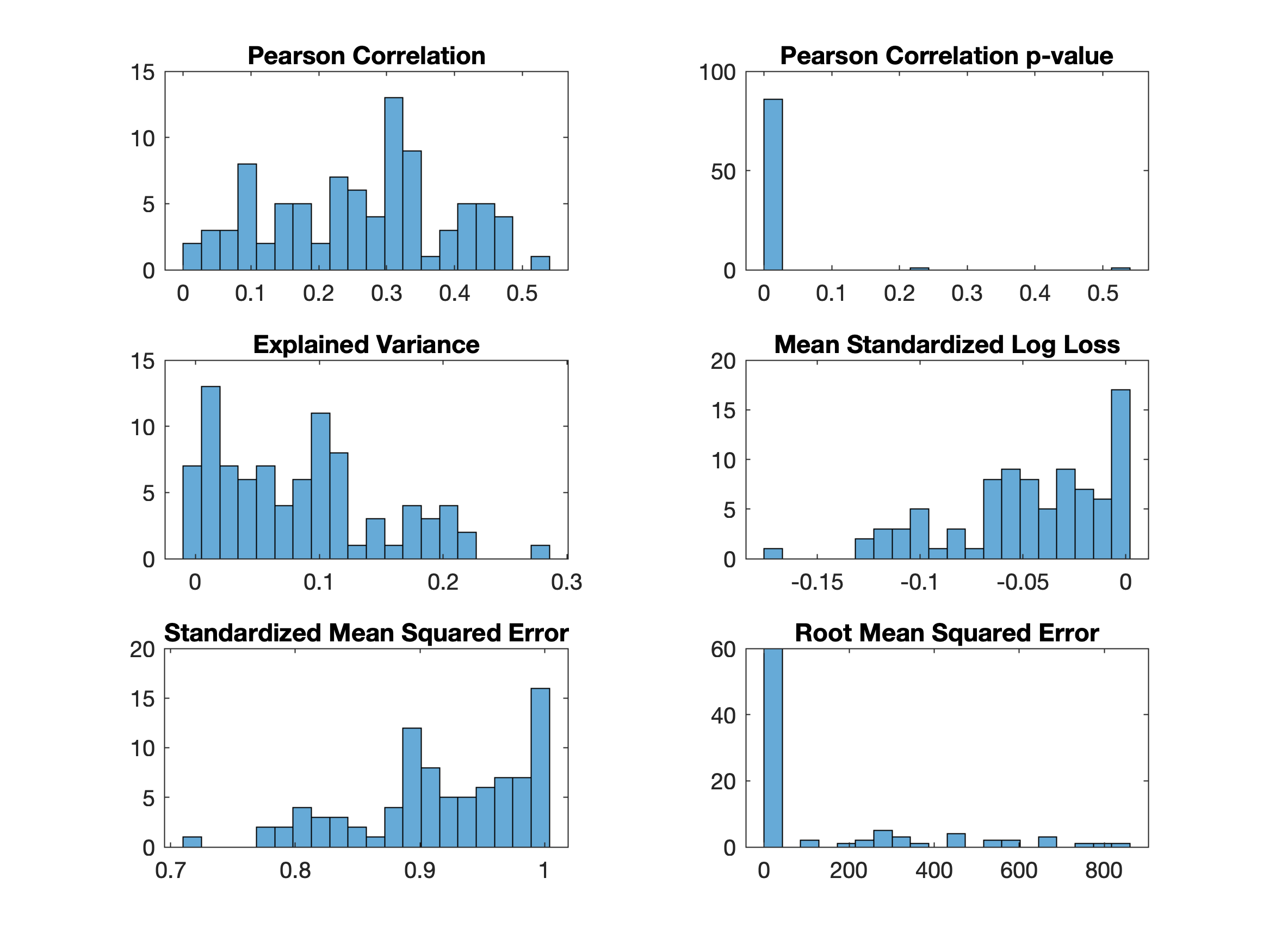

### Clinically dissociated groups supplementary comparisons

#### Longitudinal stability of clinically dissociated groups

Supplementary Figure S2. Longitudinal stability of symptom scores

To determine if the clinically dissociated groups are significantly more likely to remain higher in their defining clinical feature than the other clinically dissociated groups across other instances of RDS collection, we investigated symptom scores (including the anhedonia, depressed mood, and somatic disturbance scores) of a previous data instance (on average 6 years before the instance investigated in the main analysis) and a follow up instance (on average 2 years after the instance investigated in the main analysis). Three separate ANOVAs were performed for anhedonia, mood, and somatic scores with a factor for instance (3 levels) and a factor for clinical group (6 levels), shown here with the mean score for that clinical measure (from RDS questionnaire) for each clinically dissociated group shown on the x-axis in each panel (i.e., mean anhedonia score in panel A). The results revealed significant main effects for clinical group in all comparisons (p<0.008) and no significant main effect of instance for any comparisons (p>0.29). Follow-up analyses showed that the Anhedonia group was significantly higher (p < 2.6e-09) in their anhedonia score than the Chronic and Late Onset groups. The Low Mood group was significantly higher in the depressed mood measure than the Chronic and Late Onset groups (p<2.9e-15). The Somatic group was significantly higher in the somatic disturbance score than any other clinically dissociated group (p<0.0022) except for the acute severity group. As such, these findings show strong stability of the defining clinical features in our dissociated groups over time.

A. **Anhedonia**

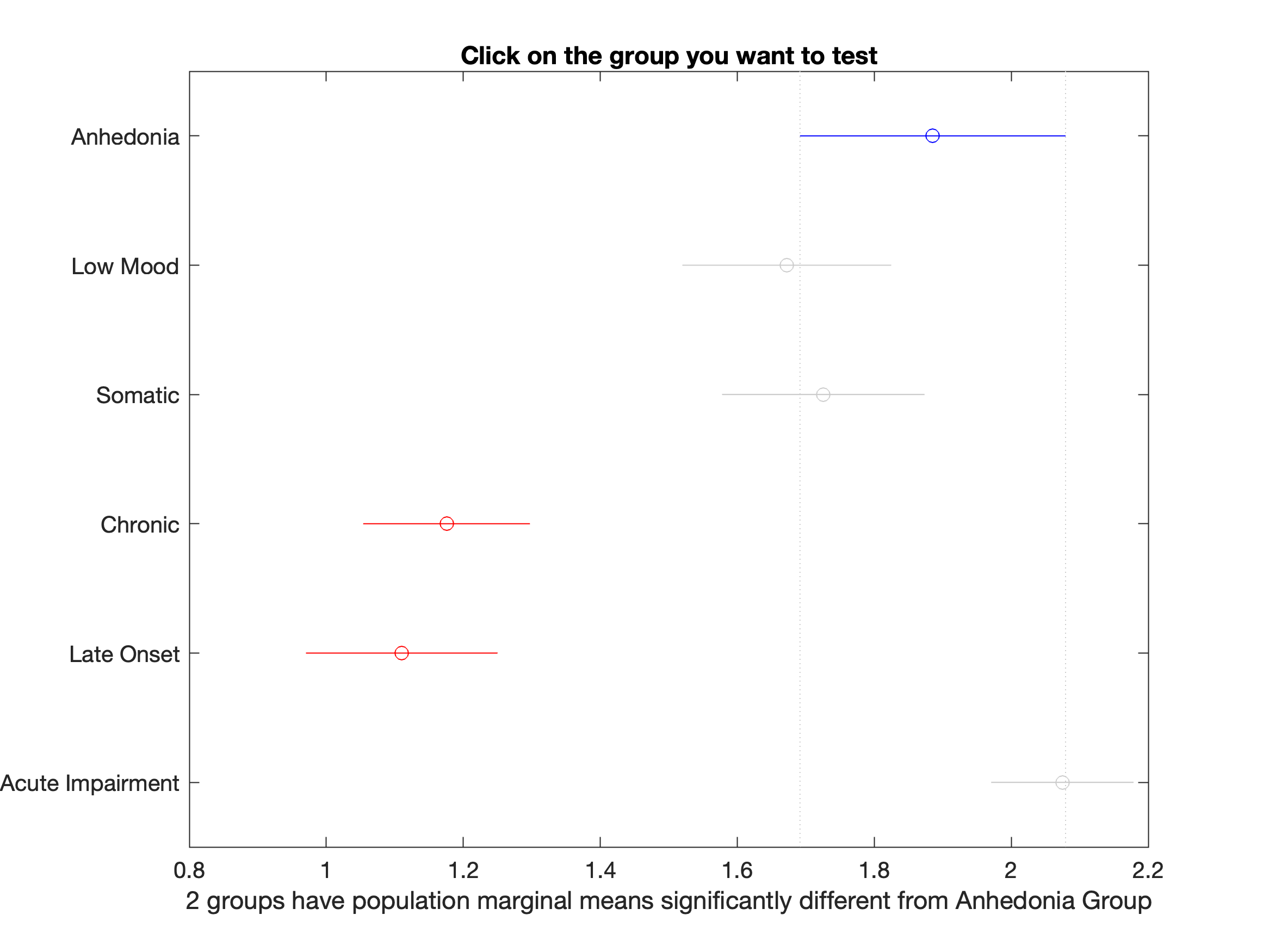

B. **Low Mood**

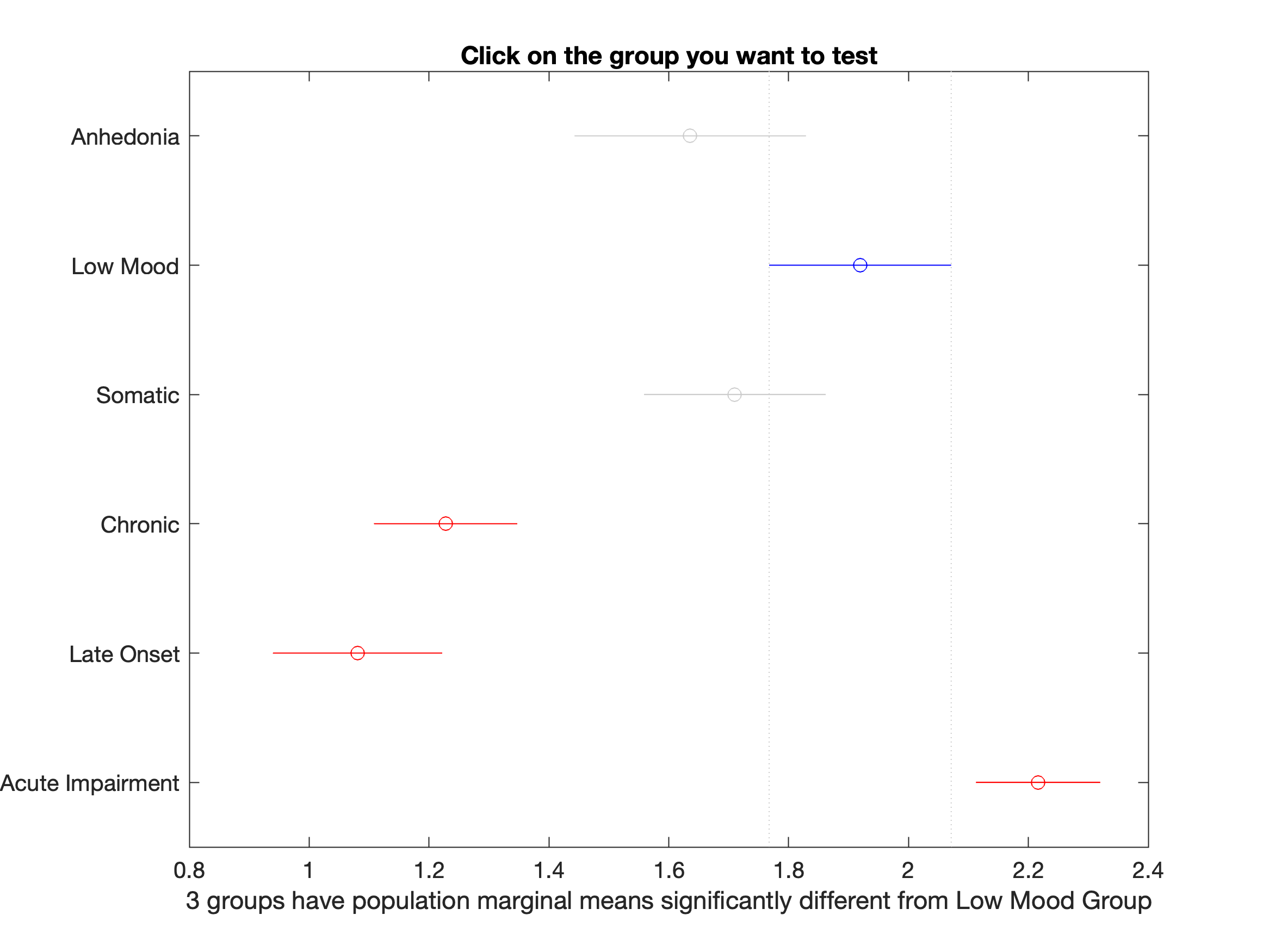

C. **Somatic**

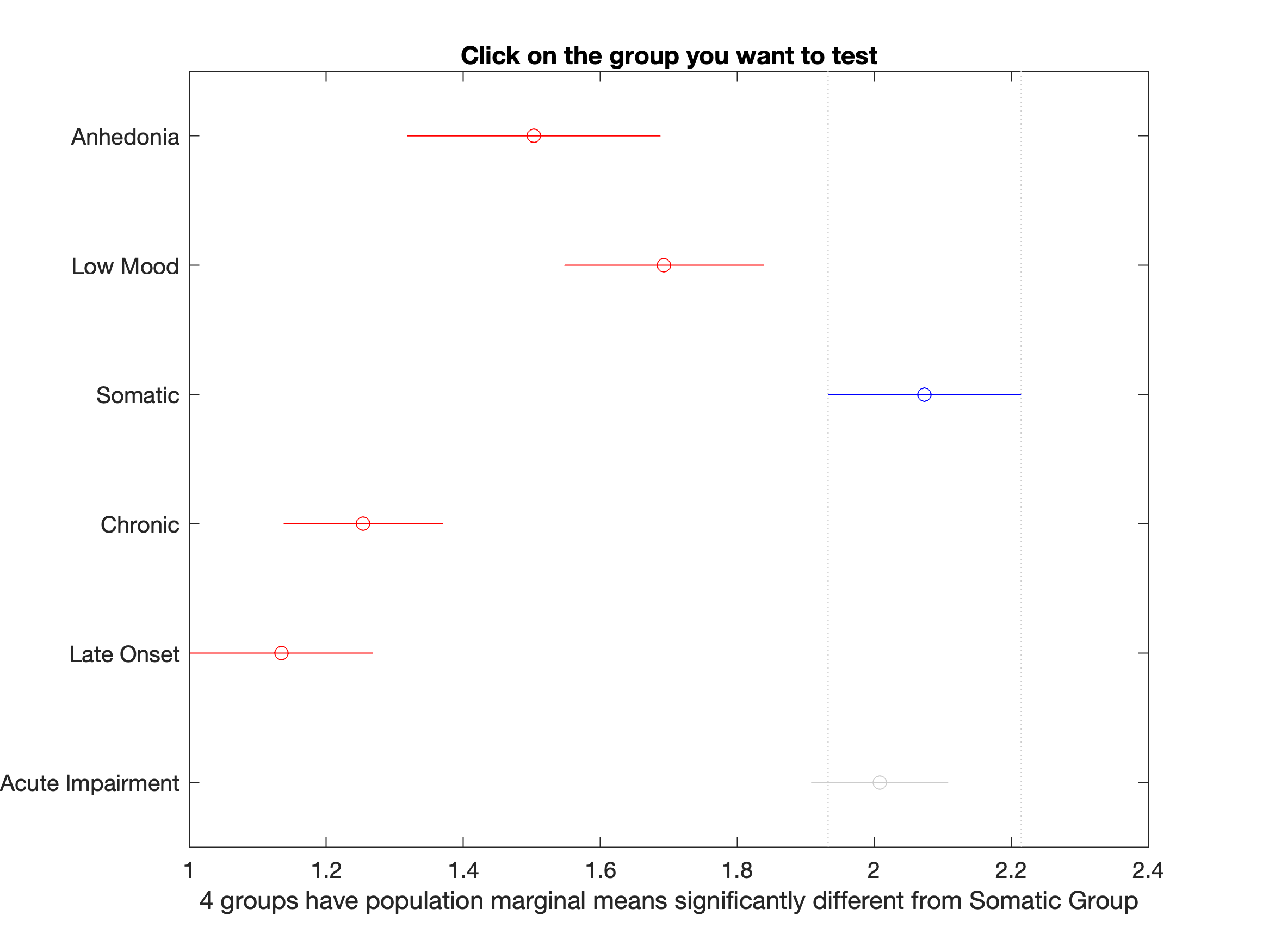

##### Supplementary Figure S3. Clinical characteristics of dissociated groups.

Mean +/- Standard Error Mean score for the clinically dissociated groups in Generalized Anxiety Disorder measure (GAD), Neuroticism measure (N), and sum Recent Depressive Symptoms (RDS Sum). As the clinically dissociated groups were selected partly based on their RDS score, a statistical analysis should be taken with limited interpretation. GAD range = 0-21, N range = 0-12, RDS range 4-16. * indicates p < 0.05, ** indicates p < 0.01, and *** indicates p < 0.001.

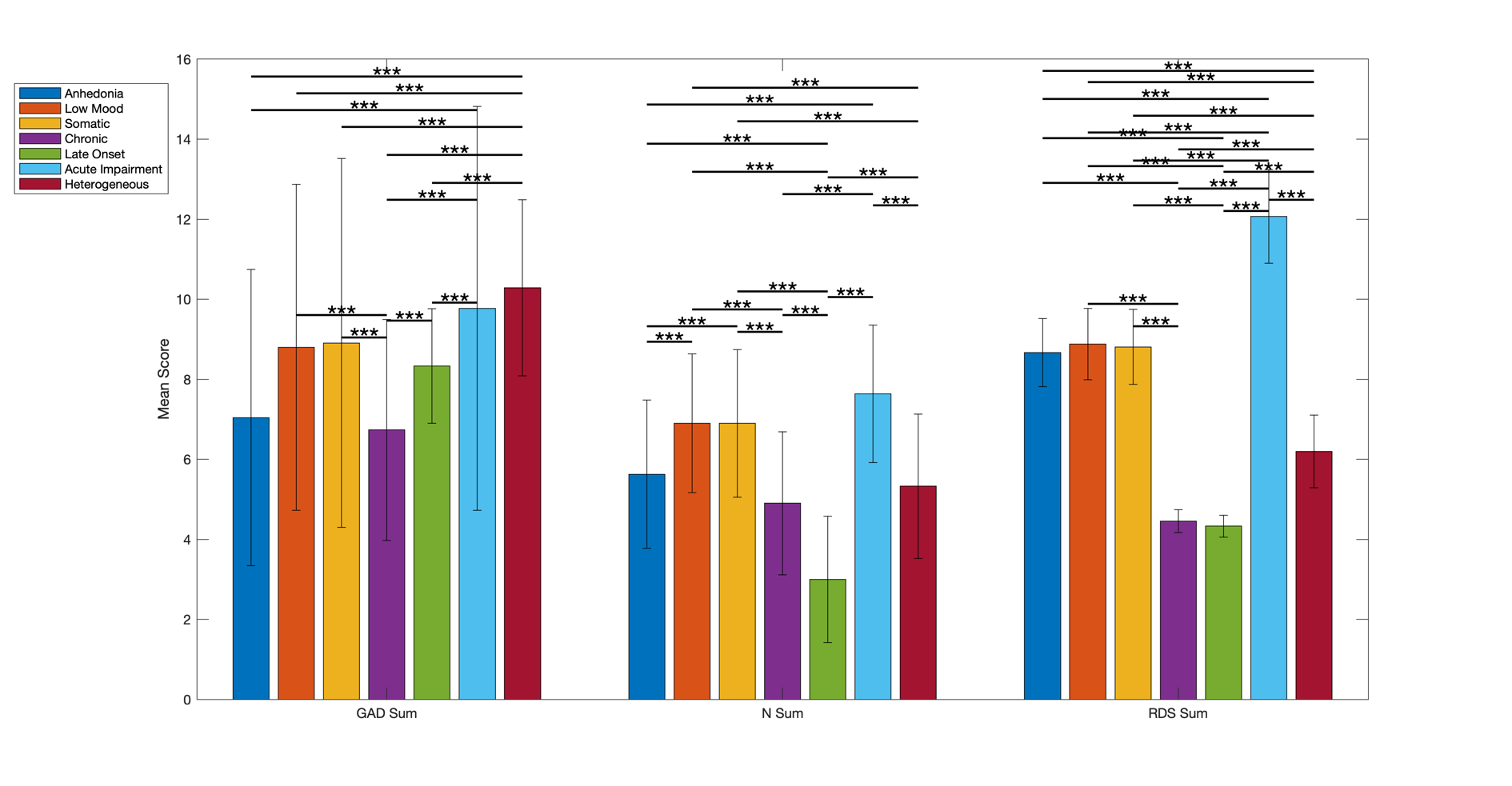

Supplementary Table S4. Differences between clinically dissociated groups.

The clinically dissociated groups were assessed for their differences in GAD (Generalized Anxiety Disorder measure) and N (Neuroticism measure). p<0.05 is bolded and the estimate of effect (with directionality) is displayed, Tukey’s multiple comparisons corrected. The estimate of effect is the estimate calculated by the Matlab ANOVA toolbox.

| Group A | Group B | GAD p | GAD estimate of effect | N p | N estimate of effect |
| --- | --- | --- | --- | --- | --- |
| Anhedonia | Low Mood | **0.004** | -1.755 | **3.37E-04** | -1.273 |
| Anhedonia | Somatic | **0.001** | -1.862 | **2.66E-04** | -1.271 |
| Anhedonia | Chronic | 0.992 | 0.308 | 0.104 | 0.725 |
| Anhedonia | Late Onset | 0.055 | -1.289 | **1.79E-21** | 2.629 |
| Anhedonia | Acute Impairment | **1.32E-10** | -2.732 | **6.80E-15** | -2.008 |
| Anhedonia | Heterogeneous | **3.18E-20** | -3.242 | 0.815 | 0.300 |
| Low Mood | Somatic | 1.00 | -0.107 | 1.00 | 0.002 |
| Low Mood | Chronic | **0.000** | 2.063 | **6.03E-13** | 1.998 |
| Low Mood | Late Onset | 0.935 | 0.466 | **0.00** | 3.902 |
| Low Mood | Acute Impairment | 0.153 | -0.977 | **0.039** | -0.735 |
| Low Mood | Heterogeneous | **2.12E-04** | -1.488 | **1.20E-12** | 1.573 |
| Somatic | Chronic | **0.000** | 2.170 | **2.11E-13** | 1.995 |
| Somatic | Late Onset | 0.829 | 0.573 | **0.00** | 3.899 |
| Somatic | Acute Impairment | 0.250 | -0.870 | **0.031** | -0.737 |
| Somatic | Heterogeneous | **5.17E-04** | -1.381 | **2.25E-13** | 1.571 |
| Chronic | Late Onset | **0.001** | -1.596 | **4.24E-14** | 1.904 |
| Chronic | Acute Impairment | **7.31E-19** | -3.040 | **0.00E+00** | -2.733 |
| Chronic | Heterogeneous | **0.00E+00** | -3.550 | 0.18 | -0.425 |
| Late Onset | Acute Impairment | **0.001** | -1.443 | **0.00** | -4.637 |
| Late Onset | Heterogeneous | **3.61E-10** | -1.954 | **0.00** | -2.329 |
| Acute Impairment | Heterogeneous | 0.234 | -0.511 | **0.00** | 2.308 |

##### Supplementary Figure S4. T-tests against 0 for all depressed participants**.**

A two-sided t-test against 0 for the normative deviations of each imaging feature (90 features) was performed on all depressed participants together. False discovery rate multiple comparisons correction was for the 90 t-tests and all 36 significant imaging features are shown here.

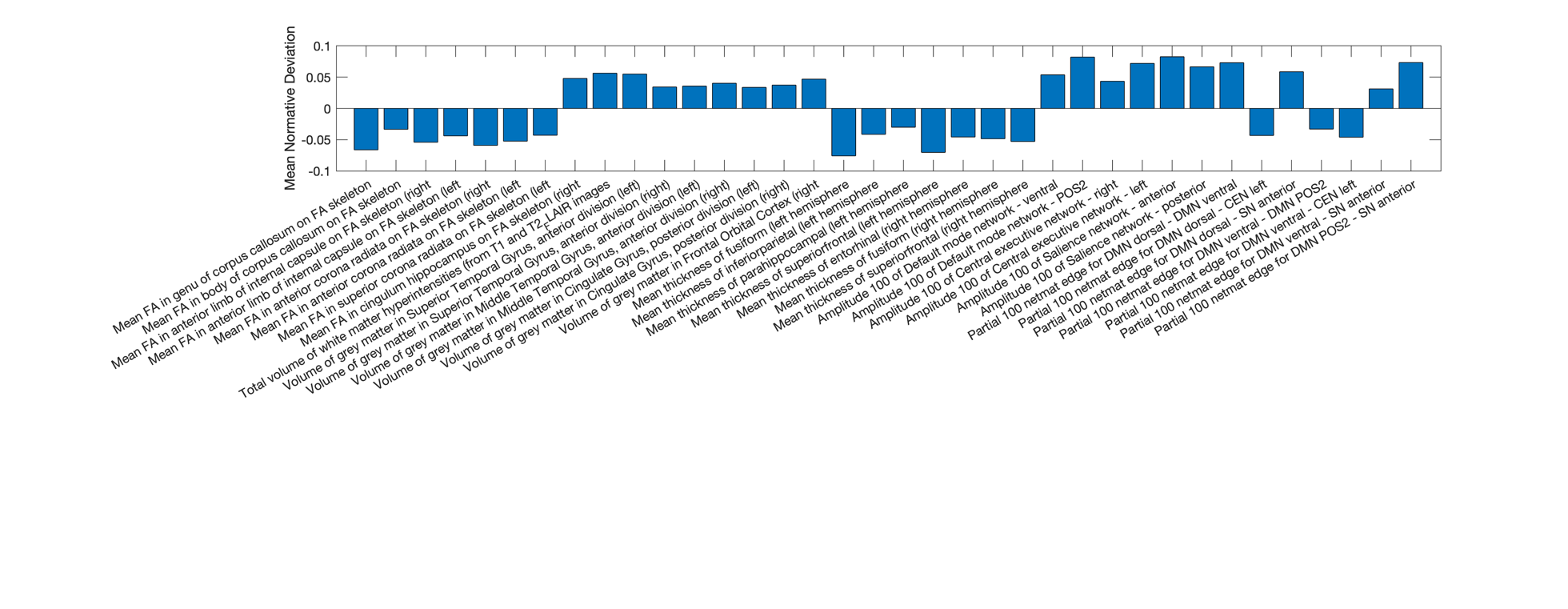

### ANOVA findings supplementary comparisons

#### Post-hoc ANOVA results

Supplementary Table S5. ANOVA Post-Hoc Results.

Multiple comparison corrected Tukey’s post-hoc differences. Only significant results are shown.

| Imaging Feature | Significant p-values |
| --- | --- |
| Superior Longitudinal fasciculus FA (R) | Depressed Mood/Late Onset p = 0.0464  Depressed Mood/Heterogeneous p = 0.0162 |
| Insula GMV (L) | Acute Impairment/Heterogeneous p = 0.0112 |
| Posterior Cingulate Gyrus GMV (L) | Depressed Mood/Somatic p = 0.0152  Depressed Mood/Acute Impairment p = 0.0478  Somatic/Heterogeneous p = 0.0058  Acute Impairment/Heterogeneous p = 0.0046 |
| Posterior Cingulate Gyrus GMV (R) | Somatic/Heterogeneous p = 0.0250  Acute Impairment/Heterogeneous p = 0.0341 |
| Frontal Orbital Cortex GMV (L) | Somatic/Heterogeneous p = 0.0244  Acute Impairment/Heterogeneous p = 0.0035 |
| Amygdala GMV (L) | Late Onset/Acute Impairment p = 0.0376  Acute Impairment/Heterogeneous p = 0.0106 |
| Amygdala GMV (R) | Somatic/Late Onset p = 0.0320  Late Onset/Acute Impairment p = 0.0336  Acute Impairment/Heterogeneous p = 0.0272 |
| Precentral Gyrus GMV (L) | Late Onset/Acute Impairment p = 0.0290  Acute Impairment/Heterogeneous p = 0.0077 |
| Precentral Gyrus GMV (R) | Somatic/Heterogeneous p = 0.0302  Acute Impairment/Heterogeneous p = 0.0145 |
| Superiorfrontal CT (L) | Somatic/Late Onset p = 0.0387  Late Onset/Acute Impairment p = 0.0013  Late Onset/Heterogeneous p = 0.0226 |

#### ANOVA comparisons against random subgroups

Supplementary Figure S4. Imaging results of clinically dissociated groups compared against random groupings.

The ANOVA was repeated on random subgroupings of sizes equal to the clinically dissociated groups over 1000 bootstraps. The resulting F-statistics are displayed as histograms for each imaging feature that was significantly different in the original ANOVA. The F-statistic for the true clinically dissociated groups (shown in red) was greater than the top 2.5% (shown in black) for every single imaging feature, most even in the top 0.5% suggesting the clinically dissociated groups are more sensitive than random subgrouping of participants.

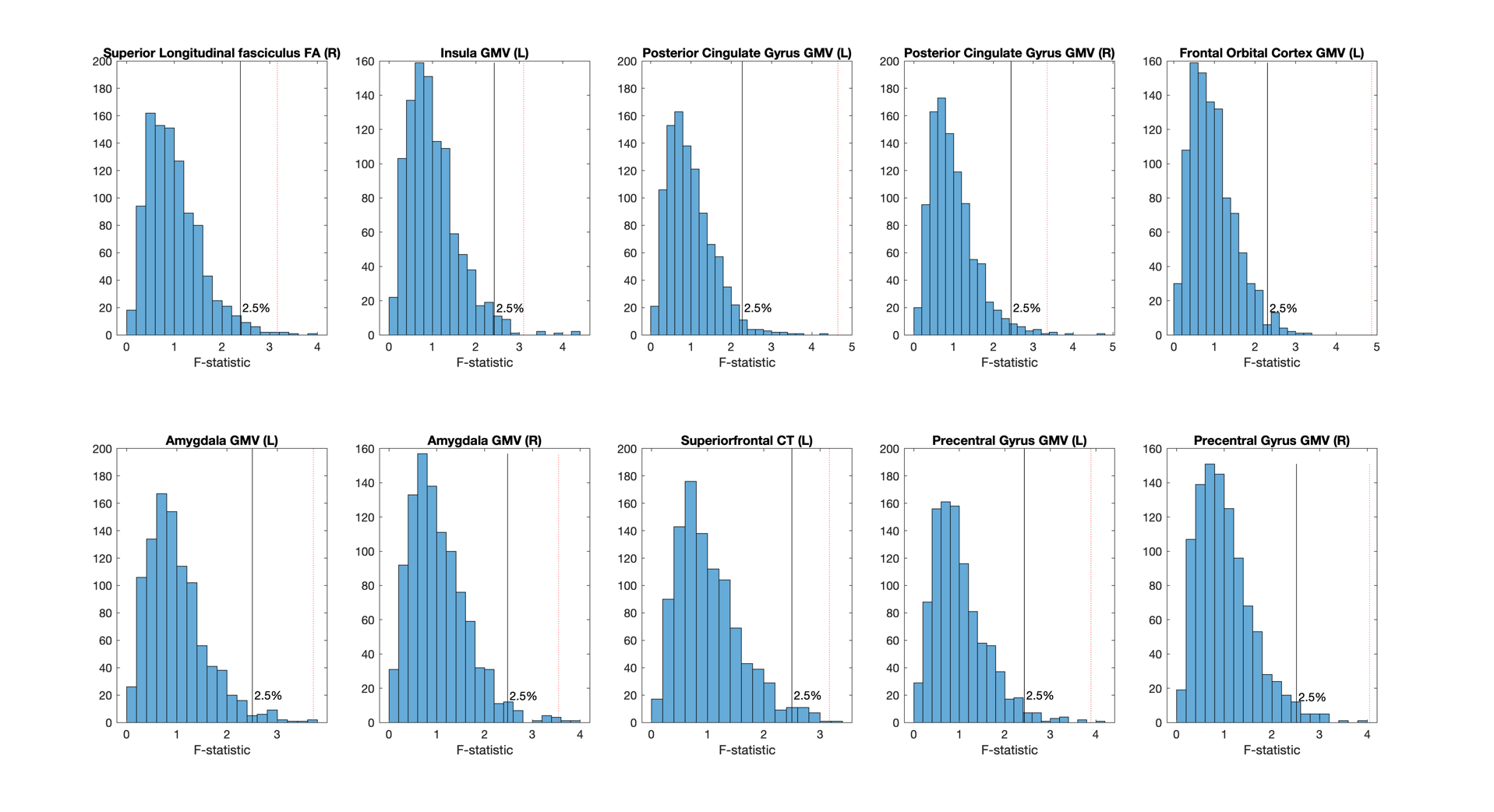

#### Reducing the heterogeneous group sample size

Supplementary Figure S5. Imaging results are not driven by heterogeneous group sample size.

The heterogeneous comparison group was substantially larger than the clinically dissociated groups, which could affect statistical significance. To assess this, the heterogeneous group was randomly subsampled at n=300 (the average size of the clinically dissociated groups) bootstrapped 1000 times. The normative deviations for the heterogeneous subsamples are displayed in the histograms with the normative deviations of the six clinically dissociated groups as the blue vertical lines. The top and bottom 2.5^th^ percentiles of the normative deviations for the heterogeneous group are visualized in red. Results confirm that the heterogeneous group had smaller neuroimaging effect sizes than one or more of the clinically dissociated groups (i.e., one or more blue lines are outside of the red lines) in 9 out of the 10 imaging features. FA = Fractional Anisotropy, GMV = gray matter volume, CT = cortical thickness.

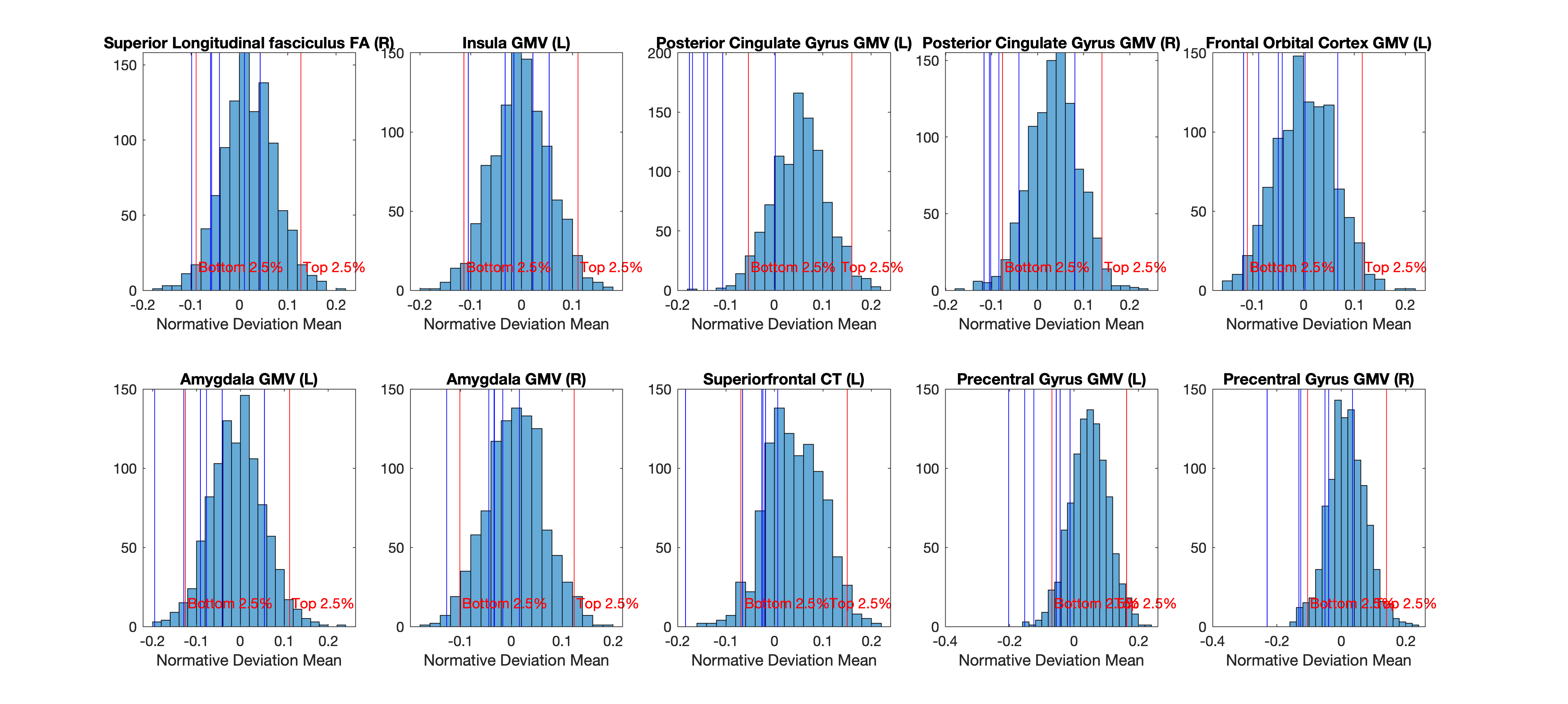

Supplementary Figure S6. Held out replication of ANOVA results.

New subjects, acquired since the original download of the UK Biobank dataset utilized for the main analyses in this study, were sorted into clinically dissociated groups (total n= 137) and heterogeneous group (total n = 296) using the same criteria. While the clinically dissociated group sizes ranged from 11 to 38, too small for a true independent replication of the main analyses, we did determine how often the normative deviations of the heterogeneous group are smaller than the normative deviations of the clinically dissociated groups over 1000 bootstraps. The heterogeneous group had smaller neuroimaging effect sizes than one or more of the clinically dissociated groups (i.e., one or more blue lines are outside of the red lines) in all 10 imaging features that were significant in the main analysis. The top and bottom 2.5^th^ percentiles of the normative deviations for the heterogeneous group are visualized in red. FA = Fractional Anisotropy, GMV = gray matter volume, CT = cortical thickness.

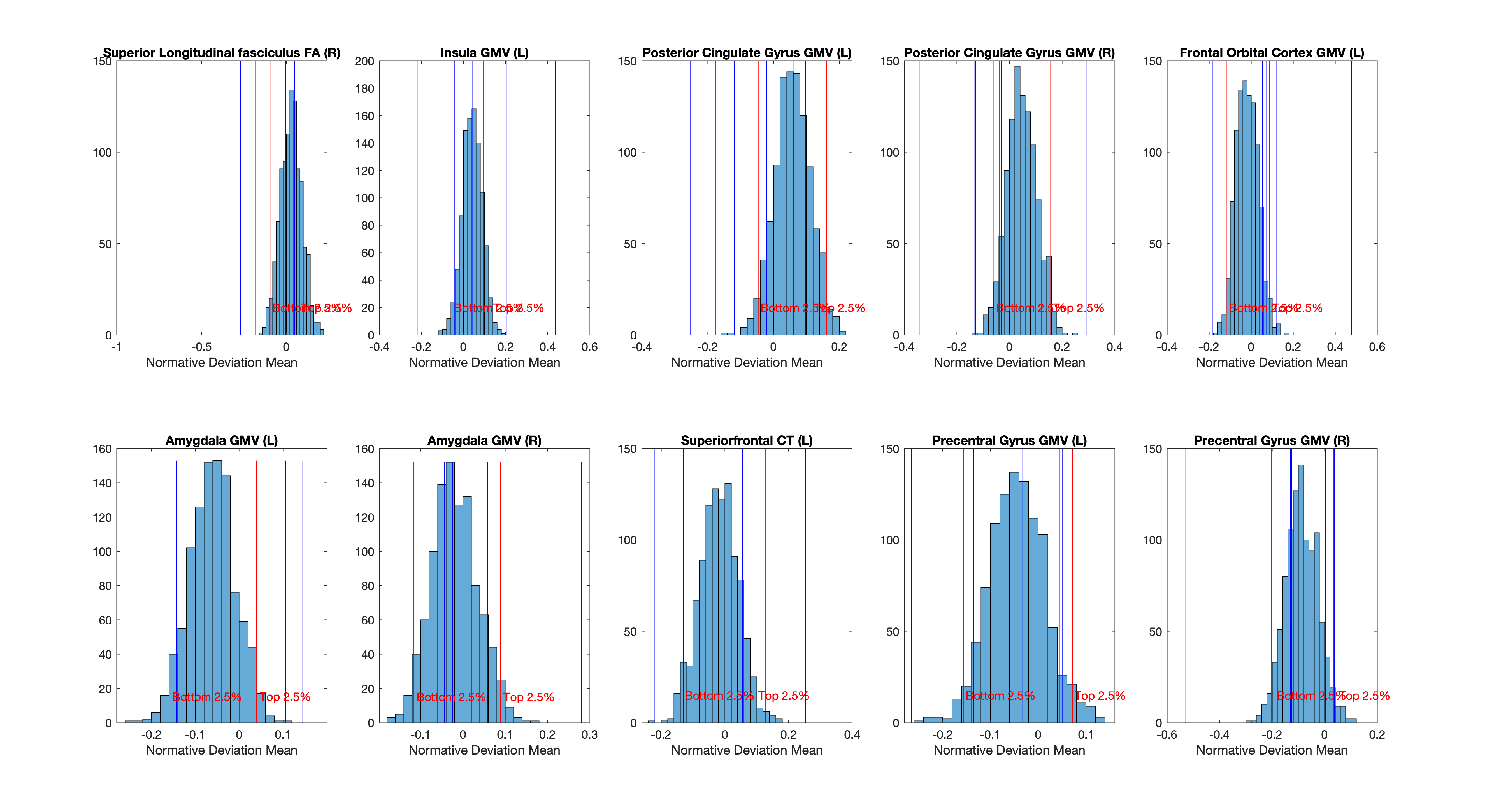

#### Held Out Sample

##### Supplementary Table S6. Characteristics of Held Out Sample.

Means if not otherwise stated. RDS = Recent Depressive Symptoms measure (sum range 4-16), individual RDS questions range 1-4.

|  | Sample Size | Age (mean±SD) | Sex | RDS Sum | RDS Anhedonia | RDS Mood | RDS Restless | RDS Lethargy | Age Onset | Episode Number |
| --- | --- | --- | --- | --- | --- | --- | --- | --- | --- | --- |
|  |  |  | (% male) |  |  |  |  |  |  |  |
| Anhedonia | 15 | 64.4±8.0 | 33.3 | 8.3 | 3.4 | 1.4 | 1.2 | 2.3 | 32.2 | 2.1 |
| Low Mood | 26 | 66.8±8.2 | 30.8 | 8.9 | 1.7 | 3.1 | 1.5 | 2.5 | 31.7 | 1.8 |
| Somatic | 17 | 64.1±10.0 | 47.1 | 8.9 | 1.7 | 1.6 | 3.2 | 2.4 | 20.4 | 3.2 |
| Chronic | 11 | 72.0±6.2 | 27.3 | 4.6 | 1.0 | 1.0 | 1.1 | 1.5 | 33.4 | 49.5 |
| Acute Impairment | 30 | 74.1±4.9 | 43.3 | 4.3 | 1.0 | 1.0 | 1.1 | 1.2 | 65.6 | 1.4 |
| Late Onset | 38 | 62.8±8.6 | 13.2 | 12.1 | 3.1 | 3.2 | 2.7 | 3.1 | 31.4 | 3.0 |
| Heterogenous | 296 | 64.9±8.1 | 37.5 | 6.2 | 1.3 | 1.4 | 1.4 | 2.0 | 34.6 | 7.6 |

### Clustering findings supplementary comparisons

#### Cluster imaging profiles

Supplementary Figure S7. Cluster Imaging Profiles.

Shown are the imaging features of the clusters determined for each clinically dissociated group, averaged for each of the 4 modalities. FA = fractional anisotropy; GMV = gray matter volume; CT = cortical thickness.

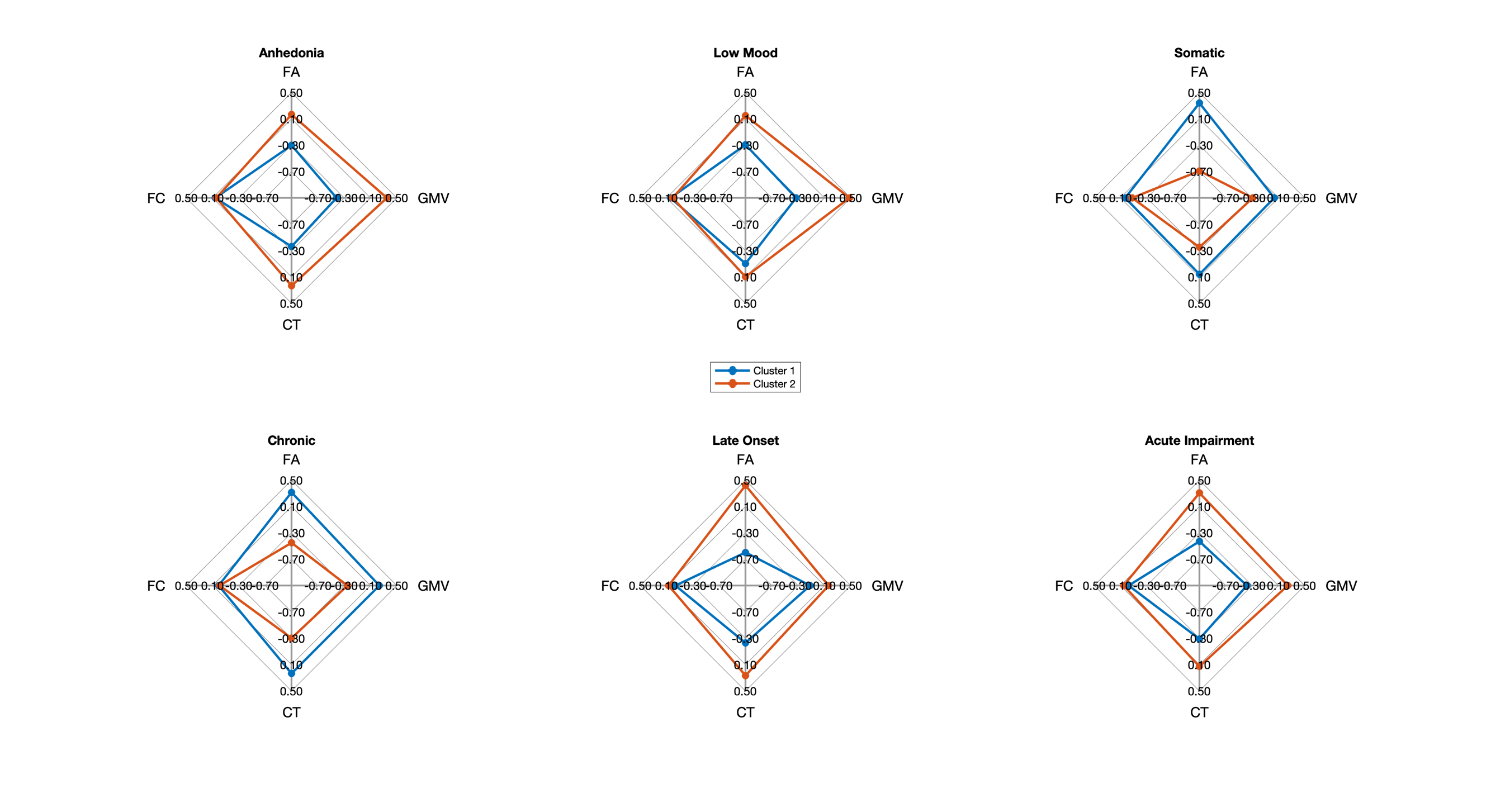

#### Clustering silhouette scores

Supplementary Figure S8. Number of clusters across bootstraps.

Shown is the optimal cluster number for each bootstrap for each clinically dissociated group. 2 was the optimal cluster number (as determined by Silhouette score) the majority of the time for each clinically dissociated group.

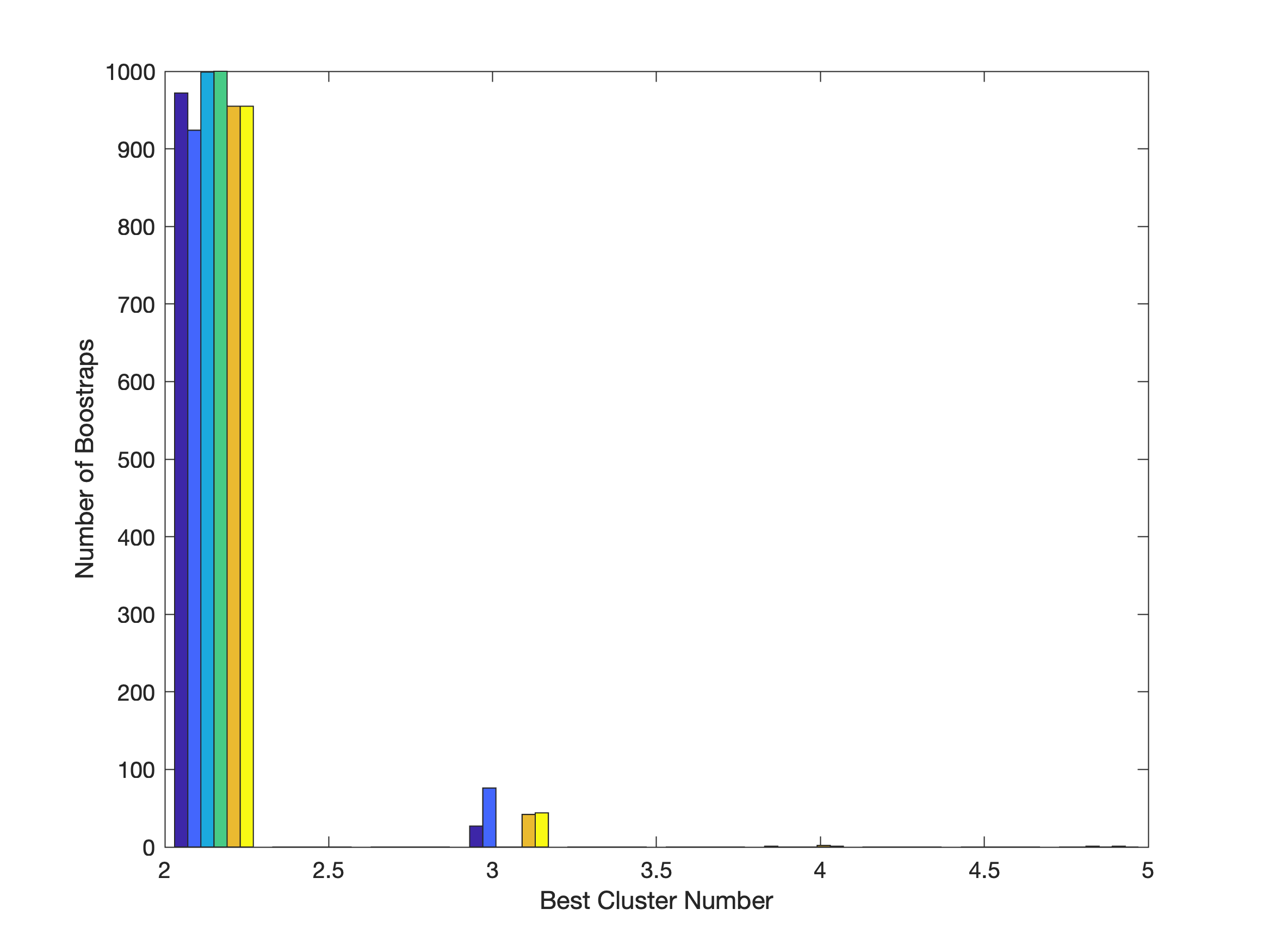

#### Clustering differentiation results

##### Supplementary Table S7. Cluster Differentiation Results.

Clusters were compared on cognition, neuroticism, and Townsend deprivation index to test their clinical differentiation. T-statistics and corrected p-values from separate unpaired t-tests in each clinically dissociated group are shown. Significant results are in bold.

|  | Cognition t | Cognition p (FDR) | Neuroticism t | Neuroticism p (FDR) | Townsend t | Townsend p (FDR) |
| --- | --- | --- | --- | --- | --- | --- |
| Anhedonia | 1.900 | 0.185 | 0.079 | 0.723 | 0.704 | 0.723 |
| Low Mood | 0.252 | 1.202 | 0.203 | 0.154 | 1.960 | 0.154 |
| Somatic | 0.135 | 1.071 | 0.105 | 1.095 | 0.110 | 1.095 |
| Chronic | -1.782 | 0.155 | -0.103 | 0.415 | -1.263 | 0.415 |
| Late Onset | -0.123 | 0.903 | -0.184 | 0.974 | 0.033 | 0.974 |
| Acute Impairment | **-3.506** | **0.003** | 0.730 | 0.079 | 2.485 | 0.079 |
